# Radial extracorporeal shock wave therapy is more effective than a combination of physical therapy modalities for rotator cuff injury: a randomized controlled trial

**DOI:** 10.1101/2025.01.07.25320065

**Authors:** Zheng Wang, Lan Tang, Ni Wang, Lihua Huang, Christoph Schmitz, Jun Zhou, Yingjie Zhao, Kang Chen, Yanhong Ma

**Author notes:** These authors contributed equally to this work (Zheng Wang, Lan Tang). **Correspondence** Dr. Yanhong Ma, Department of Rehabilitation Medicine, Shanghai Sixth People’s Hospital, Affiliated to Shanghai Jiao Tong, University School of Medicine, 600Yishan Rd, Shanghai, 200233, China.

## Abstract

**Objectives:** This study tested the hypothesis that in treatment of rotator cuff (RC) injury, radial extracorporeal shock wave therapy (rESWT) is more effective than a combination of physical therapy modalities (PTMs; i.e., interferential current therapy, shortwave diathermy and magnetothermal therapy).

**Methods:** A total of 60 patients with RC injury without presence of full-thickness RC tear were randomly allocated to rESWT for 6 weeks (n=30; one session per week) or treatment with PTMs for 6 weeks (n=30; five sessions per week). The primary outcome measure was the ASES shoulder score. Secondary outcome measures were the VAS pain score, patient’s satisfaction, shoulder range of motion, thickness of the supraspinatus tendon and the acromiohumeral distance. All outcome measures were assessed by blinded assessors at baseline as well as at 6 weeks post-baseline (W6) and W12.

**Results:** No serious adverse events occurred during the trial. Compared to the patients in the PTMs group, the patients in the rESWT group had significantly (p<0.05) higher mean ASES total scores at W6 and W12 (power with two-sided 95% confidence interval: 65.7% at W6 and 92.8% at W12) as well as lower mean VAS pain scores, higher mean satisfaction scores and higher mean active and passive shoulder abduction values at W6 and W12. Furthermore, rESWT but not PTMs significantly reduced the mean thickness of the ST and increased the acromiohumeral distance.

**Conclusion:** In treatment of rotator cuff injury without presence of full-thickness rotator cuff tear, rESWT is more effective than the investigated combination of PTMs.

**Registration:** China Clinical Trials Registration Center (Registration number: ChiCTR2300077386; registration on 7 November 2023).

## INTRODUCTION

Rotator cuff (RC) injury, including tendinitis and a tear of the RC, is one of the most common causes of shoulder pain and dysfunction, with the supraspinatus tendon (ST) being most often affected [1]. The reported incidence of RC injury varied between 13% and 37%, of which ST injury accounts for more than 90% [2,3]. Rotator cuff /ST injury can substantially affect people’s quality of life and cause profound economic and medical burden to the family and society [3].

The management of RC/ST injury includes conservative and surgical treatment modalities. According to the 2019 Clinical Practice Guidelines for the Management of Rotator Cuff Injuries of the American Academy of Orthopaedic Surgeons (AAOS), surgical repair should be considered for symptomatic, chronic, full-thickness tear as well as for acute tear and failure of physiotherapy [4]. Other RC/ST injuries, including asymptomatic, full-thickness rotator cuff tear, do not require surgery [4]. Furthermore, there is evidence that various physical therapy modalities (PTMs; including extracorporeal shock wave therapy (ESWT), interferential therapy, shortwave diathermy, magnetothermal therapy, application of ultrasound, laser, percutaneous electrical stimulation and pulsed electromagnetic field, etc.) as well as exercise rehabilitation can effectively be applied in the management of RC/ST injury [4-11]. However, randomized, controlled trials (RCTs) comparing the efficacy and safety of these PTMs in the management of RC/ST injury are largely missing.

Over the last decades, ESWT has become established as an effective and safe non-invasive treatment option for various pathologies of the musculoskeletal system [12-14]. Both focused ESWT (fESWT) and radial ESWT (rESWT) have been demonstrated to be effective and safe in the management of calcific and noncalcific tendinitis of the shoulder [15-18]. Furthermore, of particular importance was the finding in a study on rESWT for tennis elbow that partial tears of the common extensor tendon of the target elbow were healed at 24 weeks post-baseline [19]. Corresponding results have not been reported for the management of RC/ST injury with the aforementioned PTMs.

On this basis the present trial tested the hypothesis that rESWT is more effective than PTMs in the management of RC/ST injury. For obvious reasons it was impossible to test all PTMs described for RC/ST injury. Rather, a combination of interferential therapy, shortwave diathermy and magnetothermal therapy was applied in this trial.

## METHODS

### Ethics approval and study registration

This study was a prospective, randomized, controlled, single center clinical trial. It was approved by the Ethics Committee of the Shanghai Sixth People’s Hospital affiliated to Shanghai Jiao Tong University School of Medicine (No. 2023-132-(1)) in accordance with the Declaration of Helsinki, and was prospectively registered with the China Clinical Trials Registration Center (Registration number: ChiCTR2300077386). All patients signed an informed consent form before being enrolled in this trial.

### Inclusion and exclusion criteria

Female and male adults aged 18-65 years were eligible for inclusion. The inclusion criteria were: (i) unilateral shoulder pain for at least four weeks with or without limitation of shoulder range of motion (ROM), (ii) abnormal supraspinatus tendon morphology and signal intensity changes found on magnetic resonance imaging (MRI) scans enhanced with contrast agent, diagnosed as supraspinatus tendon injury or partial tear, (iii) confirmation by ultrasonography of RC/ST injury on the affected side with unaffected contralateral side, (iv) written informed consent signed and personally dated by the patient, and (v) no contraindications for rESWT and the applied PTMs.

The exclusion criteria were: (i) history of trauma of the target shoulder, (ii) bilateral shoulder pain, (iii) calcifying tendinitis of the shoulder, (iv) full-thickness tear of the supraspinatus tendon, (v) rotator cuff tear combined with other shoulder diseases such as fracture, dislocation, nerve injury, etc., (vi) local injections in the past, (vii) presence of other local diseases including active infection and active inflammatory diseases, (viii) presence of systemic diseases involving joints (e.g., rheumatoid arthritis), (ix) presence of severe osteoporosis, tumor, blood system diseases (bleeding and/or coagulation dysfunction), presence of cardiac pacemaker or defibrillator, pregnancy, sensory dysfunction, severe cognitive impairment, mental illness and epilepsy, and (x) no willingness of the patient to participate in this trial, written informed consent not signed and not personally dated by the patient.

### Sample size

The sample size was determined using the software G*Power 3.1 [20] with the following settings: test family: F tests; statistical test, MANOVA: repeated measures, within-between interaction; input parameters, two tails / effect size = 0.5 / α = 0.05 / power = 0.9 with an actual power of 0.902. Considering a potential drop-out rate of approximately 20% the final sample size was determined as n = 31 + 31 = 62.

### Randomization and Blinding

The randomization scheme was generated using the software R [21]. Sixty-nine patients were randomized with an allocation ratio of 1:1 into 11 blocks. Once a patient completed the baseline assessment, the PI retrieved the patient’s group allocation. The patient was deemed to have entered the trial at this point. No patient was replaced after randomization.

For unknown reasons three patients in the rESWT group and six patients in the PTMs group dropped out of this trial between randomization and start of the treatments (i.e., they never came back to the clinic and only baseline data of these patients were assessed). This resulted in a modified Intent-to-Treat (mITT) population of n = 30 + 30 = 60. Figure 1 shows the flow of patients through this trial according to the CONSORT statement [22].

**Figure 1.**
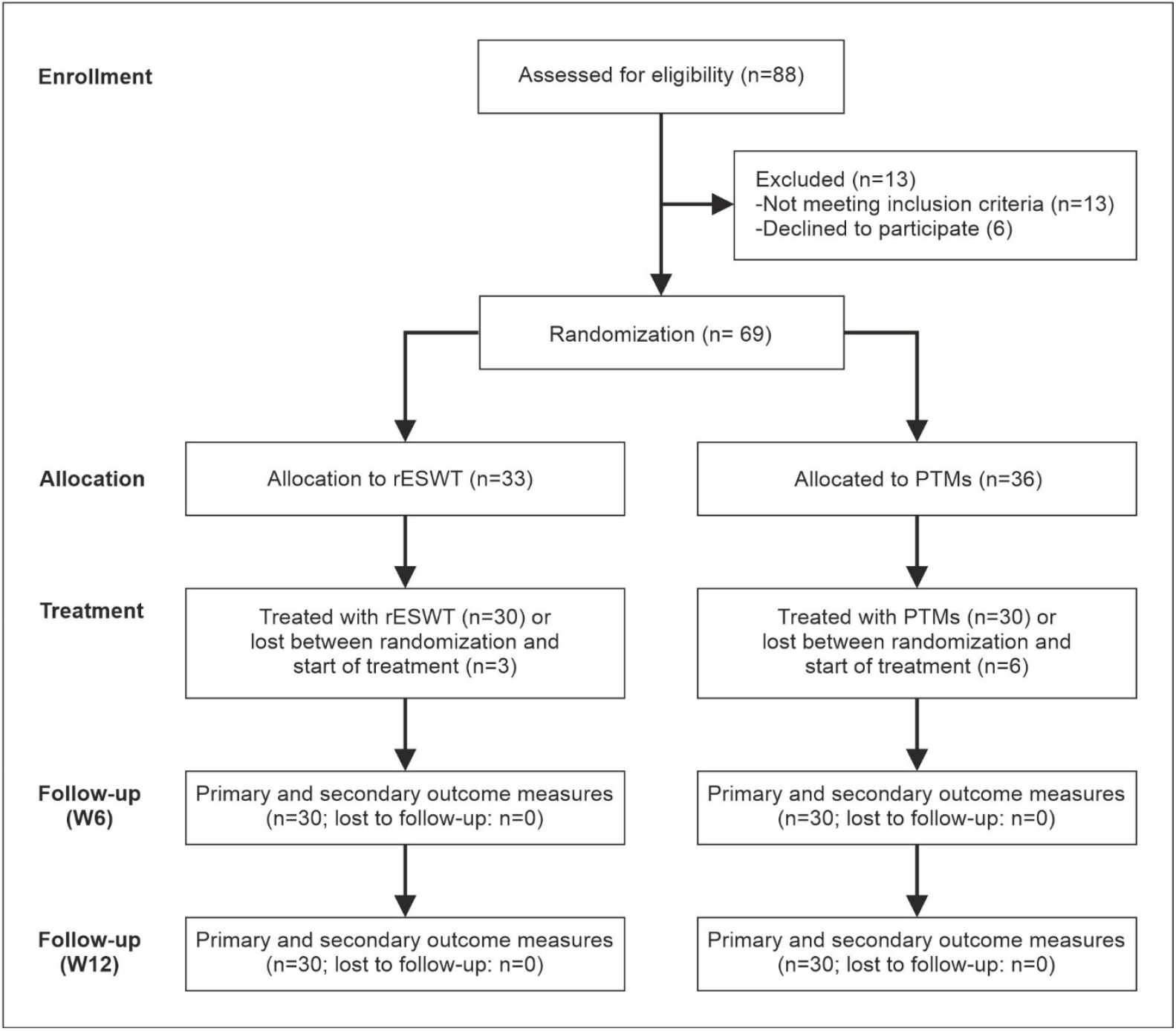
Flow of patients through this trial according to the CONSORT statement [22].

Because the patients in the two groups were treated with different treatment modalities it was not possible to blind the patients and the physiotherapists applying the treatments. On the other hand, the randomized intervention assignment was concealed from the assessors until recruitment was complete and irrevocable, and the assessors were blinded.

### Interventions

Patients in the rESWT group received the following: six rESWT sessions; one session per week; rESWT device: Swiss DolorClast (Electro Medical Systems, Nyon, Switzerland), EvoBlue handpiece, 15 mm applicator; 2000 radial extracorporeal shock waves (rESWs) per session, with positive energy flux density (EFD_+_) of 0.08 mJ/mm^2^ (achieved by operating the Swiss DolorClast at an air pressure of two bar); 8 rESWs per second, resulting in treatment time of approximately five minutes per session; application of rESWs in sitting position, with the arm of the affected side placed behind the body, the palm attached to the upper edge of the iliac spine and the fingers pointing to the pocket on the back of the pants; RC/ST injury site marked with a marker pen under the guidance of ultrasonography; application of rESWs on the marked area from various directions, centered around the target point; and no use of local anesthesia or sedative drugs.

Patients in the PTMs group received the following: 30 PTMs sessions; five sessions per week; during each session application of (i) interferential current therapy for 15 min (device: SK-10WDX, Minato Medical Science Co., Osaka, Japan), (ii) shortwave diathermy for 10 min (device: Dajia DL-C-M, Shantou Medical Equipment Factory Co., Shantou, China) and (iii) magnetothermal therapy for 20 min (device: Hot Magner HM-202 (operated in low heat mode), Chuo Medical System Co., Tokyo, Japan).

Furthermore, patients in both groups received general rehabilitation treatment, including health education and advice on how to carry out home rehabilitation exercise according to the Rotator Cuff and Shoulder Conditioning Program of the AAOS [23]. In addition, patients in both groups who had limited range of motion and needed passive joint motion used continuous passive motion (CPM; device: JKJ-1, Canwell Medical Corp., Jinhua, China) during the treatment sessions to avoid deleterious effects of immobility on the supraspinatus muscle. All patients were advised to maintain their current lifestyle during treatment and not to receive any other rehabilitation treatment.

### Primary outcome measure

The primary outcome measure was the American Shoulder and Elbow Surgeons Standardized Shoulder Assessment Form (ASES shoulder score) that shows strong correlation with multiple rotator cuff specific scores, and has excellent reliability, construct validity and responsiveness [24,25].

The ASES shoulder score consists of a pain score (relative weight: 50%) using a scale ranging from 10 (pain as bad as it can be) to 0 (no pain at all), and 10 questions (relative weight: 50%) about activities of daily life and participation in sport/leisure activities assessed on a Likert scale. The ASES total score ranges from 0 (pain as bad as it can be and poor function) to 100 (no pain at all and normal function). It has an established minimum clinically important difference (MCID) of 12-17 after conservative management of RC/ST injury [26].

The ASES total score was assessed at baseline as well as at 6 weeks post-baseline (W6) and W12.

### Secondary outcomes measures

Secondary outcome measures were the VAS Pain score (using a scale ranging from 0 (no pain at all) to 10 (pain as bad as it can be), patient’s satisfaction (using a scale ranging from 0 (maximum dissatisfaction) to 10 (maximum satisfaction)), active and passive shoulder ROM (abduction, flexion, external rotation and internal rotation, measured using an electronic goniometer (Steel Ruler Digital Angle Ruler JL-360-01, Xinliang Instrument Technology Corp., Shanghai, China), thickness of the ST and the acromiohumeral distance assessed using ultrasonography (device: SonoScape E2, SonoScape Medical Corp., Shenzhen, China).

To measure the thickness of the ST the patient was first placed in a calm state for 10 minutes. Then, the patient was placed in the same position as described above for rESWT. The ultrasonography probe was placed in front of the anterior lateral edge of the acromion, parallel to the long axis of the ST, and about 45 °between the frontal plane and the sagittal plane. The thickness of the ST was measured at the position of the most serious lesion on the affected side, as well as at the same position on the contralateratal, unaffected side.

To measure the acromiohumeral distance the patient was placed in a sitting position, with the head, shoulder, elbow joint and hand in neutral position. The ultrasonography probe was placed at the midpoint of the acromion, and the long axis of the ultrasonography probe coincided with the scapular plane of the patient. The shortest distance between the lower edge of the acromion and the surface of the upper edge of the humeral head was measured.

All secondary outcome measures were assessed at baseline, W6 and W12.

### Statistical Analysis

No patient in the mITT population was lost to follow-up. Accordingly, no missing data imputation was performed.

All outcome measures returned at each time point (baseline, W6, W12) a single data point for each patient. Normality of these data was assessed using the D’Agostino & Pearson test. Then, for each time point the group-specific mean and standard deviation (SD) were calculated. Comparison between the groups was performed using three-way repeated measures (RM) ANOVA (ST thickness) or two-way RM ANOVA (all other outcome measures) followed by Bonferroni’s multiple comparison tests.

Furthermore, relative changes of the ASES total score (RCASES), VAS pain score (RCVAS) and ST thickness (RCST) were calculated as:

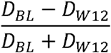

with DBL and DW12 being the corresponding, patient-specific data (ASES total score, VAS pain score or ST thickness, respectively) at baseline (BL) and at W12. Then, correlations between RCASES and RCST, RCASES and the ASES total score at baseline, RCVAS and RCST and RCVAS and the VAS pain score at baseline were calculated using linear regression analysis.

The probability value of less than 0.05 (p-value < 0.05) was considered statistically significant. In addition, the power to find a difference in the mean ASES total scores between the patients in the rESWT group and the patients in the PTMs group at W6 and W12 was calculated.

Calculations were performed using GraphPad Prism (version 10.4.1 for Windows, GraphPad Software, San Diego, CA, USA) and the software, Open Source Epidemiologic Statistics for Public Health [27].

## RESULTS

### Baseline clinical characteristics

Table 1 summarizes the clinical characteristics of the patients in the mITT population at baseline. 31 patients were male and 29 patients were female. The median / mean age of all patients in the mITT population was 41 / 41 ± 10.8 (mean ± SD) years (range, 20 – 61 years). 39 patients had RC/ST injury on the right side, and 21 patients on the left side. At baseline, the median / mean ASES total score of all patients in the mITT population was 50 / 50.8 ± 5.4 (range, 40 – 66), the median / mean VAS pain score was 5.0 / 5.2 ± 0.8 (range, 4 – 7), and the median / mean patient’s satisfaction was 4.5 / 4.4 ± 0.8 (range, 3 – 6).

**Table 1.** Baseline clinical characteristics of the patients (mITT population) enrolled in this trial. *, data given as min / median / max / mean / SD.

| Clinical characteristic | rESWT group (n=30) | PTMs group (n=30) |
| --- | --- | --- |
| Age [yr]* | 20 / 42.5 / 58 / 40.2 / 10.5 | 22 / 40.5 / 61 / 41.8 / 11.1 |
| Gender [n] (male / female / non-binary) | 17 / 13 / 0 | 14 / 16 / 0 |
| Body height [cm]* | 150 / 168.5 / 190 / 168 / 9.1 | 150 / 164.5 / 183 / 165.9 / 9.3 |
| Body weight [kg]* | 50 / 61 / 80 / 62.56 / 8.1 | 49 / 60 / 79 / 60.5 / 8.1 |
| Body mass index [kg/m2]* | 18.1 / 22.2 / 25.9 / 22.1 / 1.7 | 17.0 / 21.9 / 26.3 / 21.9 / 2.0 |
| Affected side (left / right) | 11 / 19 | 10 / 20 |
| ASES total score* | 43 / 50.5 / 64 / 51.0 / 5.3 | 40 / 49 / 66 / 50.8 / 5.6 |
| VAS pain score* | 4 / 5 / 6 / 5.2 / 0.7 | 4 / 5 / 7 / 5.2 / 0.8 |
| Patient's satisfaction* | 3 / 4.5 / 6 / 4.4 / 0.8 | 3 / 4.5 / 6 / 4.4 / 0.8 |
| Active shoulder abduction [degrees]* | 63.4 / 96.7 / 153.3 / 99.1 / 17.4 | 64.5 / 93.6 / 150.1 / 97.7 / 18.3 |
| Passive shoulder abduction [degrees]* | 84.4 / 114.3 / 166.6 / 113.7 / 15.6 | 86.5 / 112.3 / 160.2 / 113.9 / 16.8 |
| Active shoulder flexion [degrees]* | 84.0 / 109.6 / 174.4 / 117.3 / 24.5 | 61.7 / 110.0 / 171.9 / 114.5 / 24.9 |
| Passive shoulder flexion [degrees]* | 92.7 / 126.3 / 178.8 / 133.3 / 23.4 | 90.0 / 130.5 / 176.9 / 132.8 / 23.2 |
| Active shoulder external rotation [degrees]* | 20.6 / 43.3 / 61.5 / 44.1 / 8.2 | 24.0 / 41.5 / 63.6 / 43.3 / 8.8 |
| Passive shoulder external rotation [degrees]* | 36.1 / 53.9 / 73.2 / 54.2 / 8.7 | 35.6 / 52.3 / 66.3 / 52.2 / 7.8 |
| Active shoulder internal rotation [degrees]* | 15.4 / 35.4 / 51.2 / 34.9 / 7.4 | 17.4 / 32.9 / 52.4 / 34.0 / 8.4 |
| Passive shoulder internal rotation [degrees]* | 21.5 / 44.6 / 62.0 / 44.1 / 8.4 | 27.0 / 42.8 / 65.1 / 43.9 / 7.8 |
| Thickness of the supraspinatus tendon (affected side) [mm]* | 4.4 / 5.8 / 7.1 / 5.8 / 0.8 | 4.5 / 5.7 / 7.3 / 5.7 / 0.8 |
| Thickness of the supraspinatus tendon (contralateral, unaffected side) [mm]* | 3.9 / 5.3 / 6.2 / 5.2 / 0.6 | 3.7 / 5.0 / 6.8 / 5.2 / 0.7 |
| Acromiohumeral distance [mm]* | 8.3 / 10.8 / 12.4 / 10.8 / 0.9 | 9.2 / 10.9 / 13.1 / 10.8 / 0.9 |

The shoulder ROM at baseline of all patients in the mITT population was at follows (all data provided as median / mean ± SD / range [degrees]): active abduction, 94.9 / 98.4 ± 17.7 / 63.4 – 153.3; passive abduction, 113.2 / 113.8 ± 16.1 / 84.4 – 166.6; active flexion, 109.7 / 115.9 ± 24.5 / 61.7 – 174.4; passive flexion, 127.7 / 133.1 ± 23.1 / 90.0 – 178.8; active external rotation, 42.5 / 43.7 ± 8.5 / 20.6 – 63.6;

### ASES total score

The ASES total score data of both groups in the mITT population at each time point (baseline, W6, W12) passed the D’Agostino & Pearson test for normal distribution (all p values >0.05).

Both rESWT and treatment with PTMs significantly increased the mean ASES total score from baseline to W6 and further to W12 (rESWT: from 51.0 ± 5.3 (mean ± SD) at baseline to 67.7 ± 6.6 at W6 and 81.5 ± 5.4 at W12; treatment with PTMs: from 50.6 ± 5.6 at baseline to 63.7 ± 6.3 at W6 and 76.1 ± 6.9 at W12), with significantly better outcome after rESWT than after treatment with PTMs at W6 and W12 (Fig. 2A and Table 2).

**Table 2.**
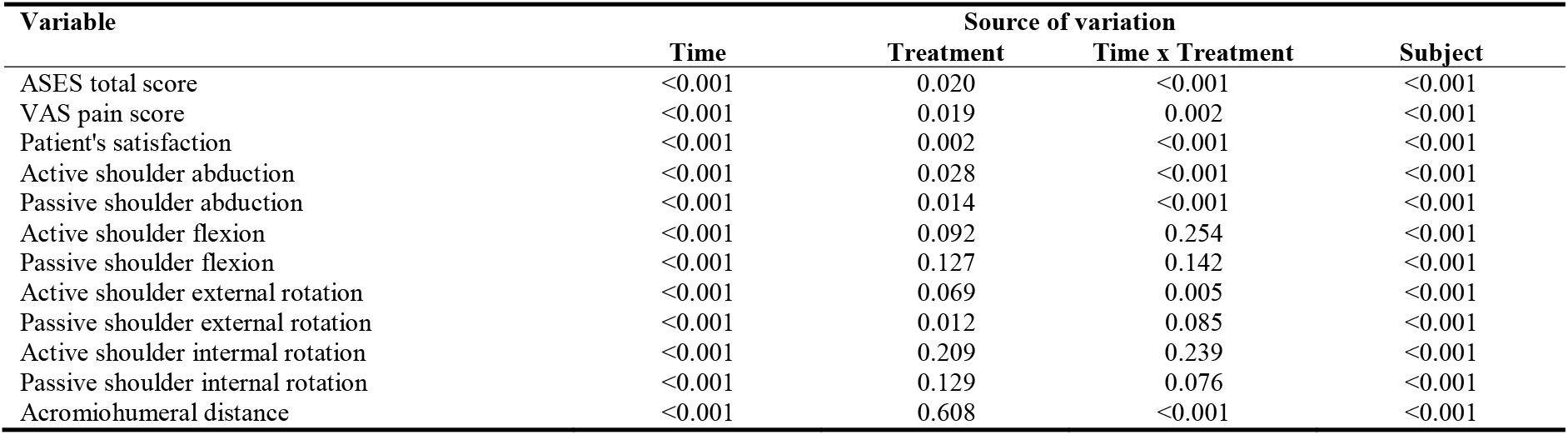
Results (p values) of the statistical analysis of the results of this trial (except of the thickness of the supraspinatus tendon) using two-way repeated measures ANOVA. The results of Bonferroni’s multiple comparison tests (rESWT vs treatment with PTMs at baseline as well as at six weeks post-baseline (W6) and W12) are shown in Figs 2, 4 and 5.

**Figure 2.**
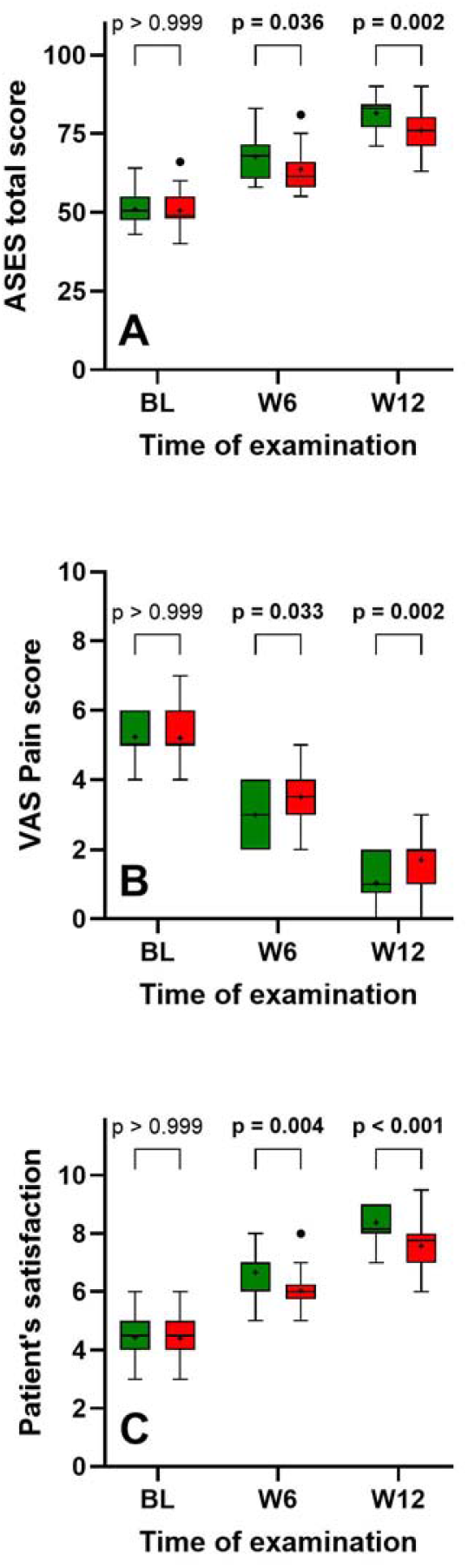
Tukey boxplots of the ASES total score (**A**), VAS pain score (**B**) and patient’s satisfaction (**C**) of the patients in the rESWT group (green bars) and the patients in the PTMs group (red bars) at baseline (BL) as well as at six weeks post-baseline (W6) and W12. The results of Bonferroni’s multiple comparison tests (rESWT vs treatment with PTMs at BL, W6 and W12) are indicated; the results of the statistical analysis of these data using two-way repeated measures ANOVA are summarized in Table 2.

The individual RCASES data showed a significant negative correlation with the individual ASES total score at baseline, but not the individual RCST data (Fig. 3A,B).

**Figure 3.**
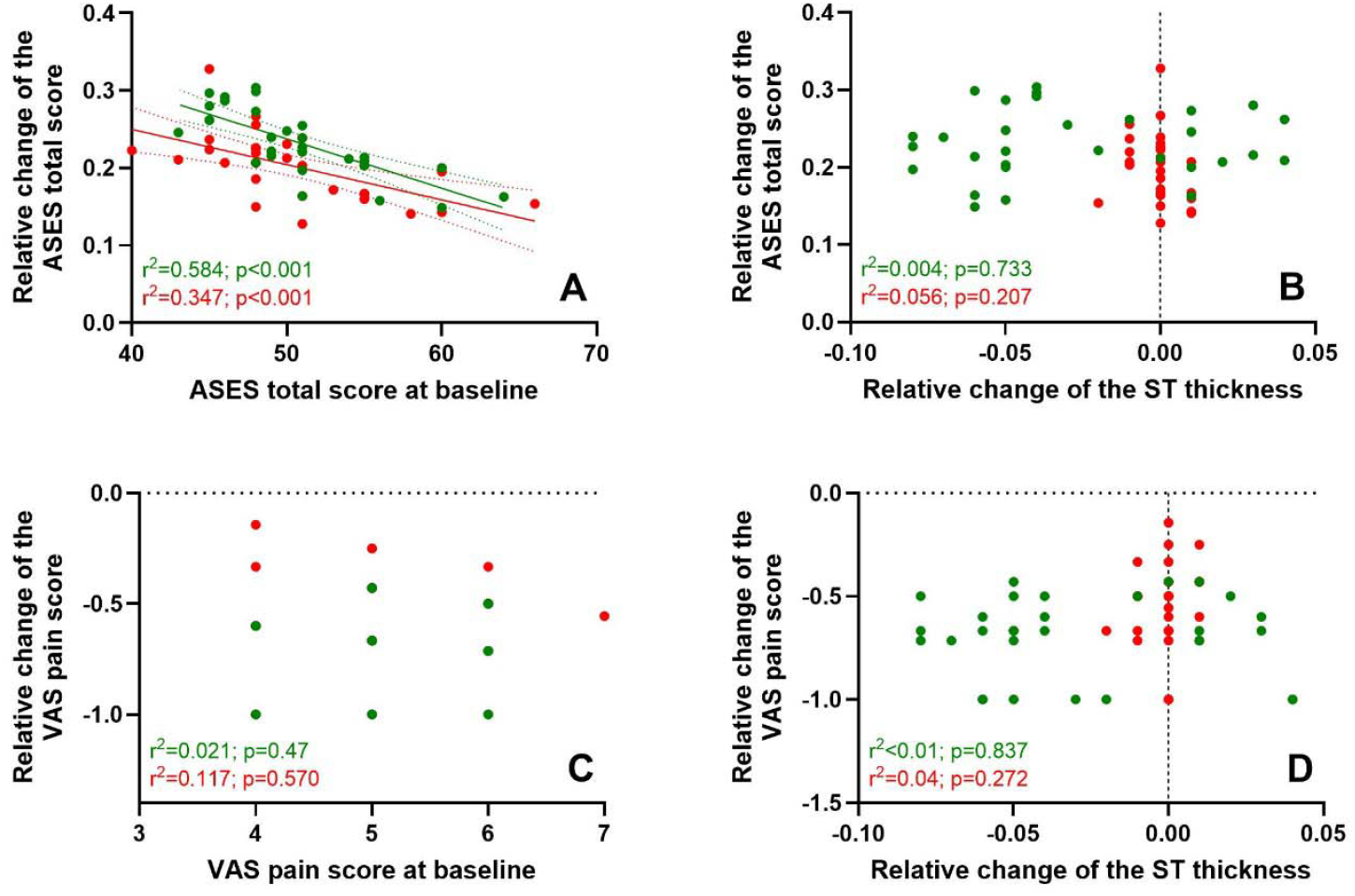
Relative change of the ASES total score from baseline to 12 weeks post-baseline (W12) as a function of the ASES total score at baseline (**A**) and the relative change of the thickness of the supraspinatus tendon from baseline to W12 (**B**), as well as relative change of the VAS pain score from baseline to W12 as a function of the VAS pain score at baseline (**C**) and the relative change of the thickness of the supraspinatus tendon from baseline to W12 (**D**) of the patients in the rESWT group (green dots) and the patients in the PTMs group (red). The results of linear regression analysis (correlation coefficient; p value of the test whether the slope of the linear regression line was significantly non-zero) are provided in the lower left corner of each panel. In case of p<0.05 the linear regression lines and their 95% confidence bands are shown.

With a two-sided 95% confidence interval this trial had a power of 65.7% at W6 and 92.8% at W12 to find a difference between rESWT and treatment with PTMs in the management of RT/ST injury. Thus, the null hypothesis had to be rejected.passive external rotation, 53.1 degrees / 53.2 ± 8.2 / 35.6 – 73.2; active internal rotation, 35.3 / 34.5 ± 7.9 / 15.4 -52.4; and passive internal rotation, 43.4 / 44.0 ± 8.1 / 21.5 – 65.1.

The median / mean ST thickness on the affected side at baseline of all patients in the mITT population was 5.7 mm / 5.7 ± 0.8 mm (range, 4.4 mm – 7.3 mm), and 5.2 mm / 5.2 ± 0.6 mm (range, 3.7 mm – 6.8 mm) on the contralateral, unaffected side. Furthermore, the median / mean acromiohumeral distance at baseline of all patients in the mITT population was 11.0 mm / 10.9 ± 0.8 mm (range, 9.0 – 13.1 mm).

### VAS pain score and patient’s satisfaction

The VAS pain score and patient’s satisfaction data of both groups in the mITT population at each time point (baseline, W6, W12) passed the D’Agostino & Pearson test for normal distribution (all p values >0.05).

Both rESWT and treatment with PTMs significantly reduced the mean VAS pain score from baseline to W12 (rESWT: from 5.2 ± 0.7 to 1.0 ± 0.7; treatment with PTMs: from 5.2 ± 0.8 to 1.7 ± 0.8) and improved the mean patient’s satisfaction from baseline to W12 (rESWT: from 4.4 ± 0.8 to 8.4 ± 0.6; treatment with PTMs: from 4.4 ± 0.9 to 7.6 ± 0.7). As in case of the mean ASES total score, rESWT resulted in significantly better outcome than treatment with PTMs at W6 and W12 (Fig. 3B,C and Table 2).

The individual RCVAS data showed no significant correlation with the individual VAS pain score at baseline or the individual RCST data (Fig. 3C,D).

### Shoulder range of motion

Except for 15 of 48 (31.3%) data sets the shoulder ROM data of both groups in the mITT population at each time point (baseline, W6, W12) passed the D’Agostino & Pearson test for normal distribution (all p values >0.05). Those data sets that did not pass the D’Agostino & Pearson test (p<0.05) are marked with asterisks in Figure 4. All of these data sets contained outliers that prevented passing the D’Agostino & Pearson test, as p values < 0.05 were obtained after removing these outliers from the corresponding data sets.

**Figure 4.**
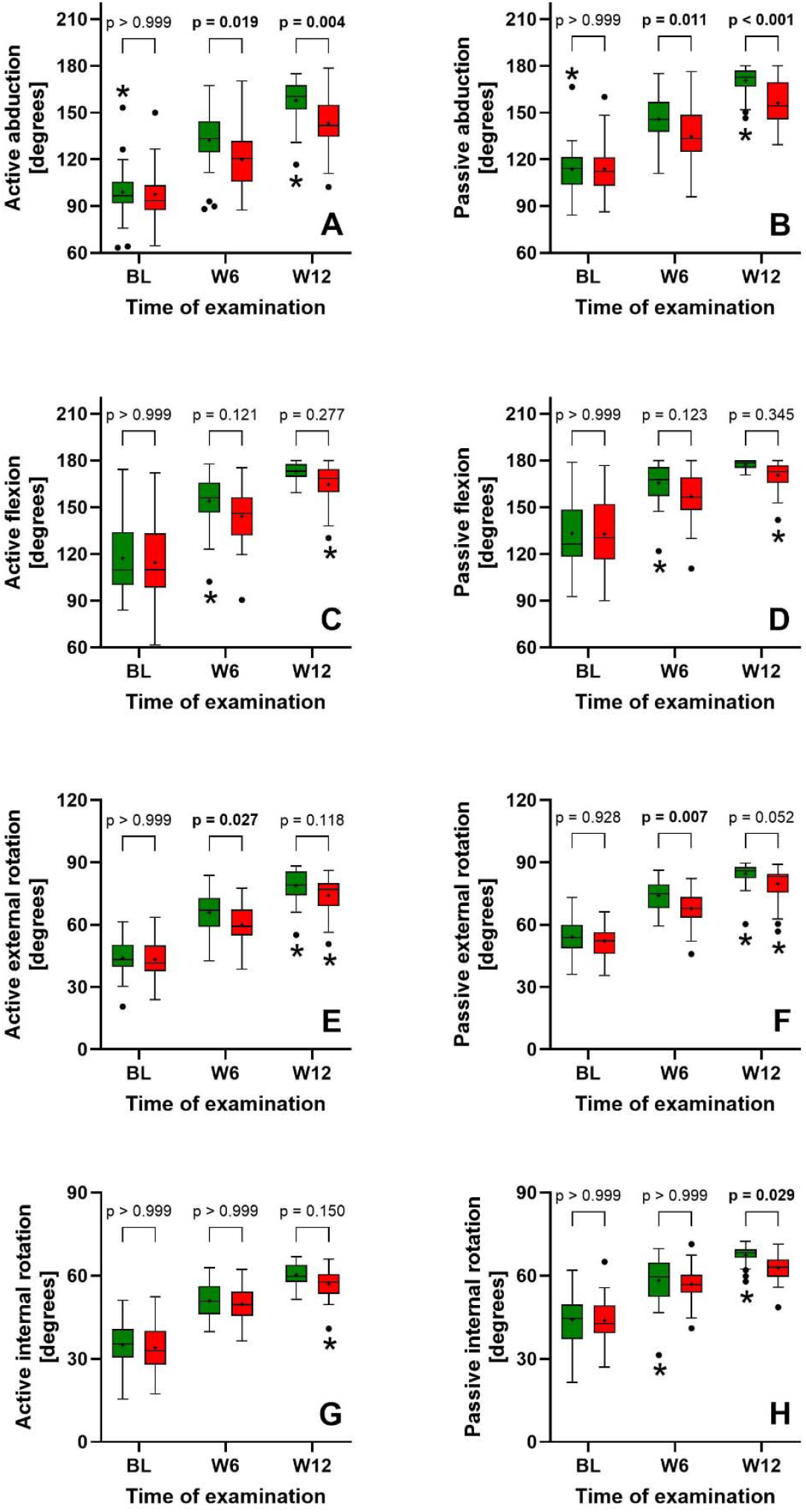
Tukey boxplots of the active and passive shoulder abduction (**A**,**B**), shoulder flexion (**C**,**D**), shoulder external rotation (**E**,**F**) and shoulder internal rotation (**G**,**H**) of the patients in the rESWT group (green bars) and the patients in the PTMs group (red bars) at baseline (BL) as well as at six weeks post-baseline (W6) and W12. The results of Bonferroni’s multiple comparison tests (rESWT vs treatment with PTMs at BL, W6 and W12) are indicated; the results of the statistical analysis of these data using two-way repeated measures ANOVA are summarized in Table 2. Those data sets that did not pass the D’Agostino & Pearson test for normal distribution are marked with asterisks.

Both rESWT and treatment with PTMs significantly increased the mean active and passive shoulder abduction from baseline to W12 (active abduction, rESWT: from 99.1 ± 17.4 degrees to 157.9 ± 13.9 degrees; treatment with PTMs: from 97.7 ± 18.3 degrees to 143. 2± 16.6 degrees; passive abduction, rESWT: from 113.7 ± 15.6 degrees to 170.8 ± 8.5 degrees; treatment with PTMs: from 113.9 ± 16.8 degrees to 156.2 ± 13.9 degrees), with significantly better outcome after rESWT than after treatment with PTMs at W6 and W12 (Fig 4A,B and Table 2).

Furthermore, both rESWT and treatment with PTMs significantly increased the mean active and passive shoulder flexion, external rotation and internal rotation. Significantly better outcome after rESWT than after treatment with PTMs was found for active and passive external rotation at W6 as well as for passive internal rotation at W12 (Fig. 4D-H and Table 2).

### Thickness of the supraspinatus tendon

Except for the patients in the rESWT group on the affected side at baseline (p=0.015) the ST thickness data of both groups in the mITT population at each time point (baseline, W6, W12) passed the D’Agostino & Pearson test for normal distribution (all p values >0.05).

Radial ESWT but not treatment with PTMs significantly reduced the mean ST thickness from baseline to W12 (rESWT: from 5.8 ± 0.8 mm to 5.4 ± 0.5 mm; treatment with PTMs: 5.7 ± 0.8 mm at baseline and at W12). There were no significantly different outcomes between the treatments at W6 and W12. On the other hand, the patients in the rESWT group had a significantly higher mean ST thickness on the affeted side than on the contralateral, unaffected side only at BL but not at W6 and W12, whereas the patients in the PTMs group had a significantly higher mean ST thickness on the affeted side than on the contralateral, unaffected side at BL as well as at W6 and W12 (Fig. 5A).

**Figure 5.**
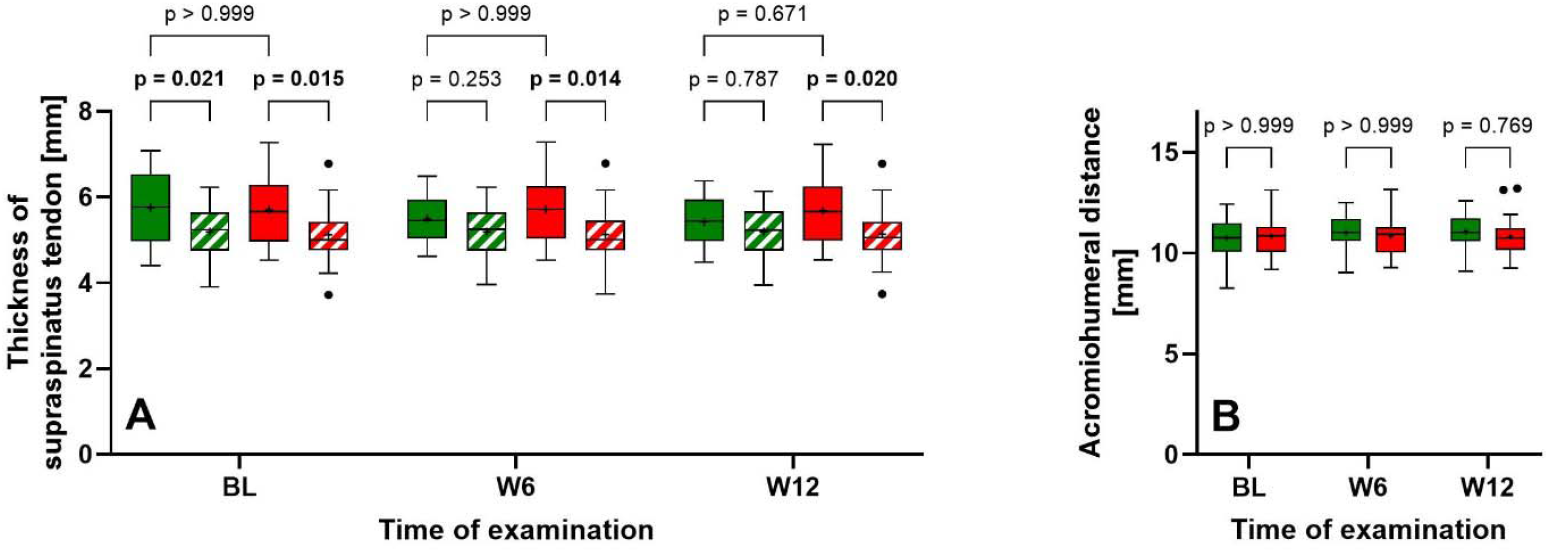
Tukey boxplots of the thickness of the supraspinatus tendon (ST thickness) (**A**) and the acromiohumeral distance (**B**) of the patients in the rESWT group (green bars) and the patients in the PTMs group (red bars) at baseline (BL) as well as at six weeks post-baseline (W6) and W12. Solid bars in (A) represent the results obtained on the affected side, and crosshatched bars the results obtained on the contralateral, unaffected side. The results of Bonferroni’s multiple comparison tests (rESWT vs treatment with PTMs as well as the ST thickness on the affected side vs the ST thickness on the contralateral, unaffected side at BL, W6 and W12) are indicated. The results of the statistical analysis of the data shown in (A) using three-way repeated measures ANOVA were as follows: time, p<0.001; treatment, p=0.781; side, p<0.001; time x treatment, p<0.001; time x side, p<0.001; treatment x side, p=0.239; and time x treatment x side, p<0.001. The results of the statistical analysis of the data shown in (B) using two-way repeated measures ANOVA are summarized in Table 2.

### Acromiohumeral distance

Except for the patients in the PTMs group at W12 (p=0.015) the acromiohumeral distance data of both groups in the mITT population at each time point (baseline, W6, W12) passed the D’Agostino & Pearson test for normal distribution (all p values >0.05).

Radial ESWT but not treatment with PTMs significantly increased the mean acromiohumeral distance from baseline to W12 (rESWT: from 10.8 ± 0.9 mm to 11.5 ± 0.8 mm; treatment with PTMs: 10.8 ± 0.9 mm at baseline and at W12). On the other hand, there was no significantly different outcome between the treatments at W6 and W12 (Fig. 5B and Table 2).

### Safety

No serious adverse events occurred during this trial.

## DISCUSSION

The results of this trial can be summarized as follows: (i) treatment of RC/ST injury with both rESWT and a combination of PTMs (interferential current therapy / shortwave diathermy / magnetothermal therapy) was effective and safe; (ii) the average MCID of the ASES total score of 14.5 reported in the literature26 was exceeded by more than double (rESWT) or more than 1.75 times (PTMs), respectively; (iii) treatment of RC/ST injury with rESWT was more effective than with PTMs; and (iv) treatment of RC/ST injury with PTMs was ‘symptom-modifying’, whereas treatment with rESWT was both ‘symptom-modifying’ and ‘structure-modifying’. No comparable studies have been published so far.

Of particular importance was the finding that six rESWT sessions with 5 min treatment time per session (total treatment time, 30 min) were more effective than 30 treatment sessions with PTMs and 45 min treatment time per session (total treatment time, 1350 min). Thus, the total treatment time was reduced by 98% when using rESWT instead of applying the investigated combination of PTMs. Together with the finding that rESWT was more effective than the investigated combination of PTMs, this reduction in total treatment time makes rESWT very attractive for both patients and therapists in the management of RC/ST injury.

The results obtained with rESWT in the management of RC/ST injury in this trial join a long list of studies in which effective pain reduction and improved function were reported after rESWT for tendinopathies and other pathologies of the musculoskeletal system [12,28,29]. Furthermore, several studies on plantar fasciopathy reported a reduction in the mean thickness of the plantar fascia (measured on ultrasound images) after treatment with rESWT [30,31] or fESWT [32,33] (note that in [30,31], the same rESWT device was used that was also used in the present trial). In line with these findings the efficacy of rESWT in the management of RC/ST injury may be explained by the following findings in the literature: (i) neurogenic inflammation contributes to the pathology of rotator cuff tendinopathy [34]; (ii) ESWT is thought to play a role in the blockade of neurogenic inflammation [35]; (iii) exposure of tendons with rESWs stimulates tendon remodelling in tendinopathy by promoting inflammatory and catabolic processes that are associated with removing damaged matrix constituents [36]; and (iv) induction of the proliferation of fibroblasts by rESWs may ultimately induce healing [37]. However, other molecular and cellular mechanisms of rESWT described in the literature may also be involved [38].

An earlier study on fESWT for plantar fasciopathy reported that patients with thinner plantar fascia after treatment experienced less pain after treatment [39]. On the other hand, a more recent study showed that fESWT reduced the plantar fascia thickness, but the only predictive factor for functional recovery in terms of the American Orthopaedic Foot & Ankle Society (AOFAS) score was the patient’s functional status prior to treatment [40]. The results of the present trial shown in Figure 3 are in line with this finding. For the sake of completeness it should be mentioned that ultrasonography (as applied in this trial) has excellent intra-and inter-rater reliability in measuring the thickness of the supraspinatus tendon [41].

A former study on treatment of partial lesion of the rotator cuff (with lesion size in each of the three axes ≤ 11 mm) with fESWT (n=15; 3 treatment sessions, one session per week; 2000 focused extracorporeal shock waves (fESWs) per session with EFD = 0.05 mJ/mm^2^; no use of local anesthesia or sedative drugs) or therapeutic exercise reported a significant improvement of the mean ASES total score from 32.2 ± 14.2 (mean ± SD; range, 8.4 – 51.6) at baseline to 54.9 ± 21.8 (range, 28.3 – 93.3) at W12 as well as an improvement of the mean VAS pain score from 7.4 ± 1.4 (range; 5 – 10) at baseline to 3.9 ± 2.2 (range, 1 – 8) at W12 after fESWT [42]. Exercise therapy did not significantly improve the mean ASES total score (baseline, 31.6 ± 11.3; W12, 35.3 ± 12.4) or the mean VAS pain score (baseline, 7.5 ± 0.8; W12, 6.9 ± 1.2).42 Regarding the mean ASES total score and the VAS pain score at baseline the patients enrolled in the former study [42] significanty differed from the patients enrolled in the present trial (Student’s t test; p < 0.001). This may also explain why the patients enrolled in the former study [42] did not experience the treatment success that was found in the present trial but required further treatment (not described in [42]). Of note, the authors of the former study [42] reported a reduction in the mean size of the partial lesion of the rotator cuff after fESWT measured on MRI scans (longitudinal axis: from 6.3 ± 1.5 mm at baseline to 5.3 ± 2.4 mm at W12; transversal axis: from 5.2 ± 1.5 mm at baseline to 4.1 ± 2.1 mm at W12; anterior-posterior axis: from 2.9 ± 0.8 mm at baseline to 1.9 ± 0.9 mm to W12). This was not found after exercise therapy (longitudinal axis: 6.4 ± 1.9 mm at baseline and 6.2 ± 2.4 mm at W12; transversal axis: 5.5 ± 1.8 mm at baseline and 5.4 ± 3.4 mm at W12; anterior-posterior axis: 2.9 ± 0.8 mm at baseline and 2.5 ± 1.0 mm to W12) [42]. The authors of [42] explained their results by findings of an earlier in vitro study in which exposure of human adipose-derived stem cells with fESWs resulted in improved differentiation of precursor cells towards tenoblast-like cells and increased synthesis of type I collagen [43]. On the other hand, one cannot rule out that the reduction in lesion size after fESWT in the former study42 was due to a reduced ST tendon thickness (as found in the present trial), as corresponding measurements were not performed in the former study [42]. Furthermore, a recent study on treatment of symptomatic, partial-thickness rotator cuff tear (sPTRCT) with injection of fresh, uncultured, unmodified, autologous, adipose-derived regenerative cells isolated at the point of care (ADRCs) found a significant improvement of the mean ASES total score and the mean VAS pain score, but no significant change in the mean tear size, during a follow-up period of 41 months (ASES total score: 58.7 ± 5.7 (mean ± standard error of the mean) at baseline, 86.1 ± 4.9 at 24 weeks post-treatment (W24), 89.4 ± 4.9 at W52, 79.6 ± 9.9 at 33.2 ± 1.0 (mean ± SD) months post-treatment (M33) and 82.3 ± 9.4 at 40.6 ± 1.9 months post-treatment (M41); VAS pain score: 4.7 ± 0.8 at baseline, 1.9 ± 0.8 at W24, 1.1 ± 0.5 at W52, 2.3 ± 1.1 at M33 and 2.4 ± 1.0 at M41; lesion size: 58.6 ± 11.3 mm3 at baseline, 45.0 ± 6.8 mm3 at W24, 44.5 ± 10.3 mm3 at W52, 55.7 ± 14.9 mm3 at M33 and 49.3 ± 14.9 mm3 at M41) [44,45].

Moreover, the analysis of a biopsy of the supraspinatus tendon obtained at 10 weeks post-treatment (W10) of a sPTRCT with injection of ADRCs (improvement of the ASES total score from 12 at baseline to 79 at W10) demonstrated that stem cells can form new tendon tissue at a site other than the original injury [46]. In addition, in a related animal model with experimentally induced partial-thickness tear of the Achilles tendon of rabbits the injection of ADRCs resulted in the formation of new tendon tissue at the lesion side, whereas the injection of saline resulted in the formation of scar tissue at the lesion side [47]. Collectively, these data strongly support the hypothesis that in the management of tendon injury, changes in the lesion size may not correlate with changes in clinical outcome measures. On this basis no measurements of the lesion size were performed in this trial.

Interferential current therapy, shortwave diathermy and magnetothermal therapy are commonly used in the treatment of musculoskeletal pain [48-50]. However, the evidence of the effectiveness of electrotherapy modalities for rotator cuff disease is low [51]. In a study on rats it was shown that interferential current therapy is effective in reducing inflammatory pain [52]. Furthermore, shortwave diathermy primarily acts on musculoskeletal tissue by inducing heat in the deep tissue, resulting in vasodilatation and increased soft tissue elasticity, amelioration of local blood flow and reduction of muscle spasms [53]. Moreover, in a study on carrageenan (CAR)-induced hindpaw inflammation in rats, treatment with magnetotherapy resulted in a reduction of the CAR-induced inflammatory edema and attenuated oxidative stress, which might be related to magnetotherapy-induced increases in the levels of antioxidant enzymes in inflamed tissues [54]. Collectively, these effects can explain the improved mean ASES score and the reduced mean VAS pain score of the patients in the PTMs group of this trial at W6 and W12. The further improvement of the clinical outcome measures of the patients in the PTMs group between W6 and W12 demonstrate that interferential current therapy, shortwave diathermy and magnetothermal therapy initiated effects in the treated tissue that lasted beyond the treatment period itself. One can reasonably hypothesize that these lasting effects were caused by the combination of these PTMs, which to our knowledge is reported here for the first time in a controlled clinical trial.

Ultrasonography is also an established method to measure the acromiohumeral distance [55] and several studies demonstrated a positive relationship between a narrowed acromiohumeral distance and the severity grading of a supraspinatus tendon tear [56-58]. Accordingly, the increased mean acromiohumeral distance of the patients in the rESWT group at W12 in this trial can be interpreted as a further indirect sign of ST tendon tissue regeneration induced by rESWT.

## Limitations

This study has five limitations. First, determining the molecular and cellular mechanisms underlying the ‘structure-modifying’ action of rESWT in the management of RC/ST injury would have required biopsies, which was not possibe in this trial. This may become the topic of future clinical trials and investigations on animal models.

Second, only one combination of PTMs was investigated. One could hypothesize that other combinations of PTMs result in similar or even better outcome than rESWT in the management of RC/ST injury. On the other hand, there is no evidence in the literature to support this hypothesis. Furthermore, given the number of treatment sessions, the treatment time and the total cost of treatment, a different combination of PTMs would have to lead to a significant improvement in clinical outcome compared to rESWT to justify the additional effort in the management of RC/ST injury.

Third, patients with full-thickness rotator cuff tear were excluded from this trial. Accordingly, the potential value of rESWT and the investigated combination of PTMs in the management of full-thickness rotator cuff tear was not assessed.

Fourth, the follow-up period of 12 weeks post-baseline was relatively short. On the other hand, in a study on fESWT for chronic plantar fasciopathy a follow-up period of 12 weeks was considered sufficient to approve fESWT for this indication by the U.S. Food and Drug Administration [59]. Future studies should assess long-term effects of rESWT in the management of RC/ST injury.

Fifth, the results of this trial should not be generalized to mean that rESWT is generally effective and safe in the management of RC/ST injury. Rather, the conclusions of this trial are restricted to the specific rEWST protocol that was applied. In this regard it must be considered that rESWs were applied at 8 Hz. At this frequency, other rESWT devices can generate rESWs with significantly lower EFD than the rESWT device used in this trial [60]. Differences in the EFD of rESWs have significant effects on treatment outcome [61,62], thereby confirming earlier findings in the management of calcifying tendinitis of the rotator cuff using fESWT [63].

## Conclusion

In treatment of RC/ST injury without presence of full-thickness rotator cuff tear, rESWT as applied in this trial is more effective than the investigated combination of PTMs, without adverse effects. Clinicians should consider rESWT instead of treatment with PTMs in the management of RC/ST injury.

## Data Availability

The datasets used and analyzed during this trial are available from the corresponding author on reasonable request, taking into account any confidentiality.

## STATEMENTS AND DECLARATIONS

## Acknowledgements

We express our gratitude to all the patients who consented to participate in this study. We also thank everyone at the Department of Rehabilitation Medicine at Shanghai Sixth People’s Hospital for their contribution to the study operations.

## Authors’ contributions

YM, ZW and LT conceived the study and participated in the design of the study. LH and JZ performed the treatments. NW and YZ performed the ultrasonography assessments. CS performed the statistical analysis and drafted the manuscript. ZW, LT and KC helped to draft the manuscript. All authors read and approved the final manuscript.

## Ethics approval and consent to participate

Ethical approval for this study was granted by the Ethics Committee of the Shanghai Sixth People’s Hospital affiliated to Shanghai Jiao Tong University School of Medicine (No. 2023-132-(1)) in accordance with the Declaration of Helsinki. The trial was prospectively registered with the China Clinical Trials Registration Center (Registration number: ChiCTR2300077386). All patients gave written informed consent.

## Consent for publication

We have obtained consent for publication.

